# Early-life Varicella Infection and Long-term Protective Associations with Chronic Disease Risk: A UK Biobank Cohort Study *

**DOI:** 10.1101/2025.03.26.25324685

**Authors:** Bingjie Li, Chen Zhu, Zhigang Yao

## Abstract

The long-term impact of early-life varicella infection on chronic disease risk remains understudied. In this UK Biobank analysis, we found that documented early-life varicella infection was associated with significantly reduced risks of multiple chronic diseases, with the strongest protective associations for neurodegenerative conditions (Alzheimer’s disease: HR = 0.23, Parkinson’s disease: HR = 0.46). These inverse associations were consistently stronger in women, increased with age, and remarkably, appeared to outweigh the effects of socioeconomic deprivation on disease risk. Mendelian randomization analyses supported causality for several outcomes. Prior varicella infection was further associated with favorable baseline health profiles, including lower BMI, improved lung function, and reduced inflammatory markers. Our findings suggest that early-life varicella infection may confer long-term protection against chronic diseases through persistent modulation of immune and inflammatory pathways, challenging simplified views of infections as uniformly harmful and highlighting the complex, time-dependent interactions between infectious exposures and chronic disease risk across the lifespan.

---

Chronic diseases-including neurodegenerative conditions such as Alzheimer’s disease (AD) and Parkinson’s disease (PD), along with cardiovascular disease (CVD), chronic kidney disease (CKD), chronic obstructive pulmonary disease (COPD), and type 2 diabetes-represent the predominant global health challenge of the 21st century [1, 2, 3]. These conditions significantly impair quality of life, diminish functional independence, and place immense economic and social burdens on healthcare systems worldwide [4, 5]. Despite extensive research into genetic, metabolic, and lifestyle risk factors, the etiology of these conditions remains partially understood, limiting the development of effective preventive strategies and disease-modifying interventions.

Emerging evidence suggests that the conventional paradigm of infections as uniformly detrimental to health requires reconsideration. The “hygiene hypothesis,” first proposed by Strachan in 1989 [6], and its modern extension, the “old friends hypothesis” [7], posit that reduced exposure to certain microorganisms may contribute to immune dysregulation and increased susceptibility to inflammatory conditions. More recently, the concept of “trained immunity”—defined as the long-term functional reprogramming of innate immune cells after exposure to certain stimuli-has emerged as a crucial mechanism through which early-life infections can shape lifelong health trajectories [8]. This process involves substantial epigenetic modifications at promoters and enhancers of inflammatory genes, alongside metabolic rewiring of immune cells, allowing them to mount enhanced responses to subsequent challenges even after returning to a non-activated state.

Early-life Varicella (chickenpox) infection provides a unique model for exploring the lifelong effects of infections on chronic disease risks. Caused by Varicella-zoster virus (VZV), varicella establishes latency in sensory ganglia after primary infection, maintaining a persistent interaction with the host immune system [9]. Recent research reveals an intriguing paradox: while later-life VZV reactivation as herpes zoster is associated with increased risk of neurological complications [10], preventing such reactivation through vaccination appears protective against dementia [11]. However, a significant gap exists in our understanding of how primary VZV infection in childhood influences long-term health outcomes. Current literature has predominantly focused on the consequences of herpes zoster in adulthood, with limited attention to the potential trained immunity effects from early-life varicella infection. This oversight represents a critical knowledge gap, particularly considering the high seroprevalence of VZV [12] and the possibility that childhood infection may induce distinct immunological programming with far-reaching implications for inflammatory and neurodegenerative disease pathways.

In this study, we conducted a novel investigation of the association between early-life Varicella infection and risk of six major chronic diseases-Alzheimer’s disease, Parkinson’s disease, cardiovascular disease, chronic kidney disease, chronic obstructive pulmonary disease, and type 2 diabetes-utilizing data from the UK Biobank cohort (N = xx). We assessed whether childhood Varicella infection alters disease risk through immune modulation and inflammatory pathways, and investigated potential variations and interactions with sex, age, and socioeconomic gradients. Additionally, we explored the biological plausibility of these relationships by investigating associations between Varicella history and multiple physiological systems, including body composition, lung function, cardiovascular health, inflammatory markers, metabolic parameters, and immune cell distributions. Through both observational and genetic data-strengthened causal inference approaches, we seek to understand the potential causal role of early-life Varicella infection in shaping lifelong health trajectories. This comprehensive investigation of early infection and chronic disease risk challenge conventional understanding of infection-disease relationships and provide novel perspectives on the developmental origins of adult disease.

## Results

In our analysis of UK Biobank participants, 25,826 (5.1%) had a documented history of varicella infection prior to baseline assessment, while 476,408 (94.9%) had no such record. Comparing baseline characteristics between groups (**Extended Data Table 1**), we found that participants with varicella infection were significantly younger (mean age 53.5 vs. 56.7 years, *p <* 0.001), predominantly of White ethnicity (97.1% vs. 94.3%, *p <* 0.001), with no significant difference in sex distribution (*p* = 0.760). The varicella infection group demonstrated higher socioeconomic status, including higher income (38.9% vs. 24.9% earning *≥* $52, 000 annually, *p <* 0.001), higher rates of college education (44.8% vs. 31.4%, *p <* 0.001), and lower deprivation scores (median TDI −2.52 vs. −2.11, *p <* 0.001). Regarding health indicators, the varicella infection group exhibited a healthier lifestyle profile, with lower BMI (median 26.0 vs. 26.8, *p <* 0.001), lower hypertension prevalence (39.9% vs. 47.3%, *p <* 0.001), and higher non-smoking rates (60.1% vs. 53.7%, *p <* 0.001). Activity levels showed statistically significant but minor practical differences between groups (*p <* 0.001), while individuals with varicella infection were more likely to have normal sleep duration (76.8% vs. 72.7%) and less likely to have long sleep (0.9% vs. 1.9%, *p <* 0.001).s

We examined the age distribution at diagnosis for Varicella and six chronic diseases of interest (**Fig. 1a**). Varicella typically occurred early in life (median age: 7.0 years, 10th-90th percentile: 4.1-26.3 years), whereas chronic diseases manifested much later. Alzheimer’s disease showed the latest onset (median age: 76.0 years), followed by Parkinson’s disease (71.3 years) and chronic kidney disease (69.4 years). Cardiovascular disease, chronic obstructive pulmonary disease, and type 2 diabetes had slightly earlier median onset ages (62.8, 65.4, and 64.2 years, respectively).

**Fig. 1.**
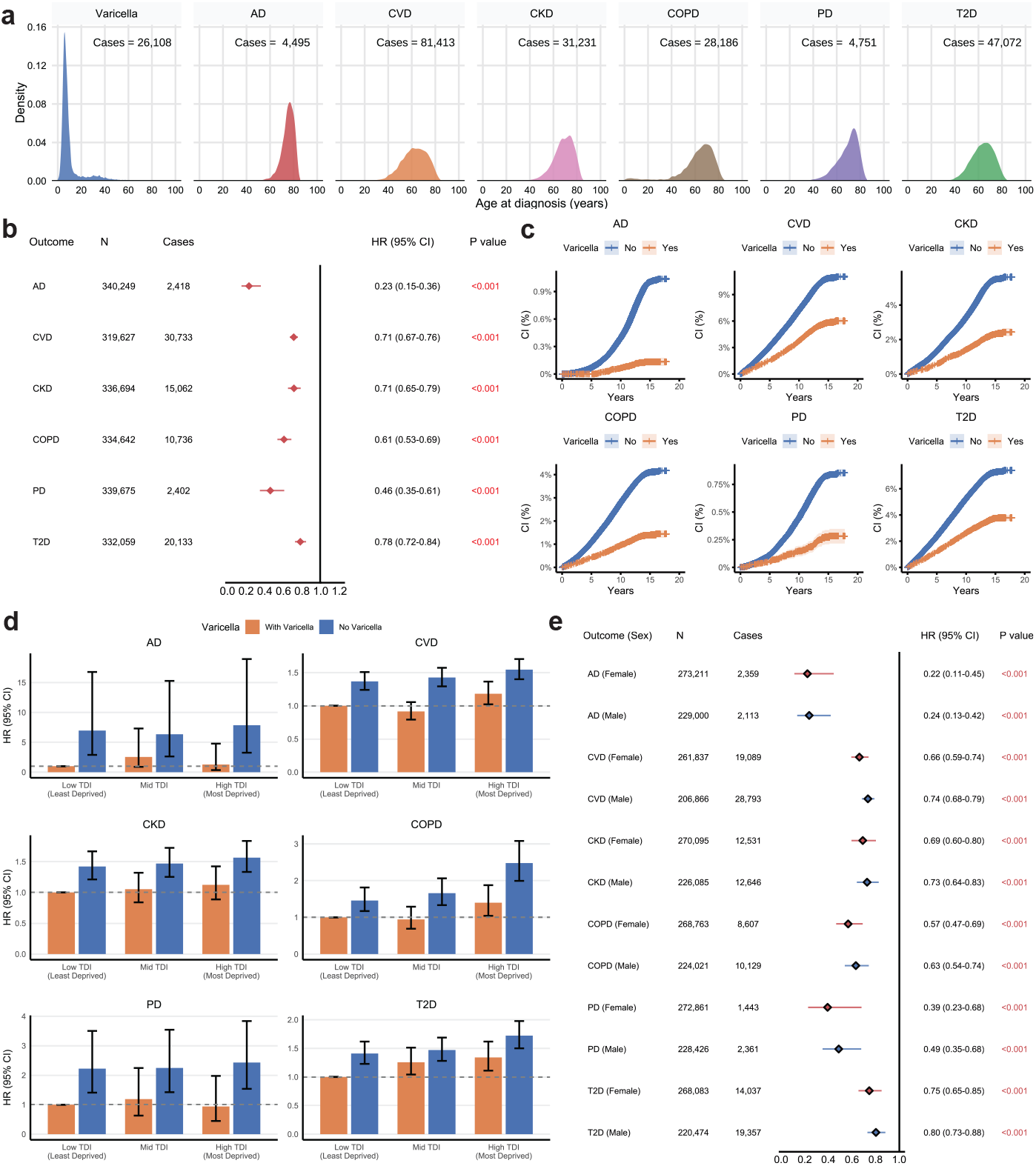
Associations between prior Varicella zoster virus (chickenpox) infection and chronic disease risk in the UK Biobank cohort. Analyses focused on six chronic diseases: Alzheimer’s Disease (AD), Cardiovascular Disease (CVD), Chronic Kidney Disease (CKD), Chronic Obstructive Pulmonary Disease (COPD), Parkinson’s Disease (PD), and Type 2 Diabetes (T2D). All Cox models were adjusted for demographics (age, sex, ethnicity, income, education, deprivation index) and lifestyle factors (smoking, sleep, BMI, physical activity). **a**. Age distribution of disease onset across the seven conditions, showing density distributions of age at diagnosis. **b**. Forest plot showing hazard ratios (HR) with 95% confidence intervals (CI) for the association between prior chickenpox infection and each chronic disease. **c**. Cumulative incidence curves comparing disease risk between participants with and without reported chickenpox infection. Shaded areas represent 95% confidence intervals. **d**. Joint effects of socioeconomic deprivation (measured by Townsend Deprivation Index tertiles) and prior chickenpox infection on chronic disease risk. The reference group (HR=1.0) is participants with low TDI and prior chickenpox infection. **e**. Sex-stratified analysis showing HRs for women and men.

Cox proportional hazards models revealed significant inverse associations between prior Varicella infection and the risk of all six chronic diseases after adjusting for sociodemographic and lifestyle factors (**Fig. 1b**). The strongest protective association was observed for Alzheimer’s disease (HR = 0.23, 95% CI: 0.15-0.36, *p <* 0.001), followed by Parkinson’s disease (HR = 0.46, 95% CI: 0.35-0.61, *p <* 0.001) and chronic obstructive pulmonary disease (HR = 0.61, 95% CI: 0.53-0.69, *p <* 0.001). Significant protective associations were also found for chronic kidney disease (HR = 0.71, 95% CI: 0.65-0.79, *p <* 0.001), cardiovascular disease (HR = 0.71, 95% CI: 0.67-0.76, *p <* 0.001), and type 2 diabetes (HR = 0.78, 95% CI: 0.72-0.84, *p <* 0.001). All associations remained highly significant after Bonferroni correction.

Cumulative incidence curves illustrated consistently lower disease incidence among participants with prior Varicella infection throughout follow-up (**Fig. 1c**). Baseline disease prevalence before prospective follow-up (**Extended Data Fig. 1**) showed higher incidence rates in the non-Varicella group across all diseases, particularly for Alzheimer’s disease (0.048 vs. 0 per 1, 000), Parkinson’s disease (1.954 vs. 0.620 per 1, 000), and cardiovascular disease (68.704 vs. 30.822 per 1, 000).

In the Mendelian Randomization analyses, varicella was further found to be causally associated with reduced risks of Alzheimer’s Disease (OR = 0.86, 95% CI: 0.80-0.92, *p <* 0.001), Cardiovascular Disease (OR = 0.98, 95% CI: 0.97-0.99, *p* = 0.002), Chronic Obstructive Pulmonary Disease (OR = 0.93, 95% CI: 0.88-0.98, *p* = 0.007), and Type 2 Diabetes (OR = 0.89, 95% CI: 0.83-0.95, *p <* 0.001) as shown in **Extended Data Fig. 2**. On the other hand, we found no evidence to suggest that the outcomes of these diseases influence the incidence of varicella, indicating the absence of bidirectional causality between these traits.

We explored the joint effects of Varicella infection and socioeconomic deprivation on chronic disease risk (**Fig. 1d**). Using individuals with low Townsend Deprivation Index (TDI) and prior Varicella infection as the reference group, we observed a striking pattern across all diseases. For Alzheimer’s disease, individuals without Varicella infection had substantially elevated risk regardless of socioeconomic status (low TDI: HR = 6.96, 95% CI: 2.89-16.78; mid TDI: HR = 6.34, 95% CI: 2.63-15.28; high TDI: HR = 7.85, 95% CI: 3.26-18.93; all *p <* 0.001). Notably, individuals with prior Varicella infection but higher deprivation did not show significantly elevated AD risk, suggesting Varicella infection may mitigate negative effects of socioeconomic disadvantage.

The magnitude of the Varicella effect notably exceeded that of socioeconomic status across multiple conditions. For example, individuals with prior Varicella infection but high socioeconomic deprivation had lower Alzheimer’s disease risk (HR = 1.28, *p* = 0.711) than individuals with no Varicella infection but favorable socioeconomic circumstances (HR = 6.96, *p <* 0.001). Similar patterns were observed for other diseases, with absence of Varicella infection consistently conferring greater risk than socioeconomic disadvantage alone. For Parkinson’s disease, having Varicella infection even with high deprivation (HR = 0.94, *p* = 0.871) was more protective than having low deprivation without Varicella infection (HR = 2.22, *p <* 0.001). For cardiovascular disease, chronic kidney disease, chronic obstructive pulmonary disease, and type 2 diabetes, we observed a gradient of increasing risk with both absence of Varicella infection and higher deprivation. The highest risk was consistently found in individuals with high deprivation and no Varicella infection, with hazard ratios ranging from 1.57 (95% CI: 1.33-1.84) for chronic kidney disease to 2.48 (95% CI: 1.99-3.08) for chronic obstructive pulmonary disease (all *p <* 0.001) compared to the reference group.

Sex-stratified analyses revealed protective associations across both sexes for all six chronic diseases, with significant sex-by-exposure interactions (all *p* for interaction *<* 0.001) (**Fig. 1e**). Notably, protective effects were consistently stronger in women across all conditions. For Alzheimer’s disease, women showed stronger protection (HR = 0.22, 95% CI: 0.11-0.45, *p <* 0.001) than men (HR = 0.24, 95% CI: 0.13-0.42, *p <* 0.001). Similar patterns were observed for Parkinson’s disease (women: HR = 0.39, 95% CI: 0.23-0.68; men: HR = 0.49, 95% CI: 0.35-0.68) and COPD (women: HR = 0.57, 95% CI: 0.47-0.69; men: HR = 0.63, 95% CI: 0.54-0.74). For cardiovascular disease, women also demonstrated stronger protection (HR = 0.66, 95% CI: 0.59-0.74) than men (HR = 0.74, 95% CI: 0.68-0.79), as did chronic kidney disease (women: HR = 0.69, 95% CI: 0.60-0.80; men: HR = 0.73, 95% CI: 0.64-0.83). The sex difference was most pronounced for type 2 diabetes, with women showing notably stronger protection (HR = 0.75, 95% CI: 0.65-0.85) compared to men (HR = 0.80, 95% CI: 0.73-0.88).

Age-stratified analyses revealed significant effect modification by age (all *p* for interaction *<* 0.001), with protective associations generally strengthening with increasing age (**Extended Data Fig. 3**). For Alzheimer’s disease, the protective effect was negligible in the youngest age group (*≤* 55 years: HR = 0.44, 95% CI: 0.14-1.41, *p* = 0.169) but became highly significant in middle-aged (55-64 years: HR = 0.15, 95% CI: 0.07-0.32, *p <* 0.001) and older adults (*>* 65 years: HR = 0.25, 95% CI: 0.14-0.47, *p <* 0.001). A similar pattern emerged for Parkinson’s disease, with no significant protection in the youngest group but strong effects in middle-aged (HR = 0.37, 95% CI: 0.25-0.56, *p <* 0.001) and older groups (HR = 0.25, 95% CI: 0.12-0.50, *p <* 0.001). For chronic kidney disease and type 2 diabetes, the protective association strengthened with age, with the strongest protection in older adults (*>* 65 years: HR = 0.57, 95% CI: 0.46-0.70 and HR = 0.64, 95% CI: 0.52-0.80, respectively, both *p <* 0.001).

Beyond disease outcomes, we examined the relationship between prior Varicella infection and baseline health parameters. Among physical measures (**Fig. 2a**), Varicella infection was most strongly associated with lower BMI (*β* = −0.088, *p <* 0.001) and body fat percentage (*β* = −0.064, *p <* 0.001). Significant positive associations were observed with multiple lung function parameters, including FEV1 (*β* = 0.064, *p <* 0.001), FVC (*β* = 0.061, *p <* 0.001), and FEV1 predicted percentage (*β* = 0.091, *p <* 0.001). Varicella infection was also associated with greater muscular strength (grip strength: *β* = 0.043, *p <* 0.001) and favorable cardiovascular parameters, including lower systolic (*β* = −0.054, *p <* 0.001) and diastolic blood pressure (*β* = −0.044, *p <* 0.001).

**Fig. 2.**
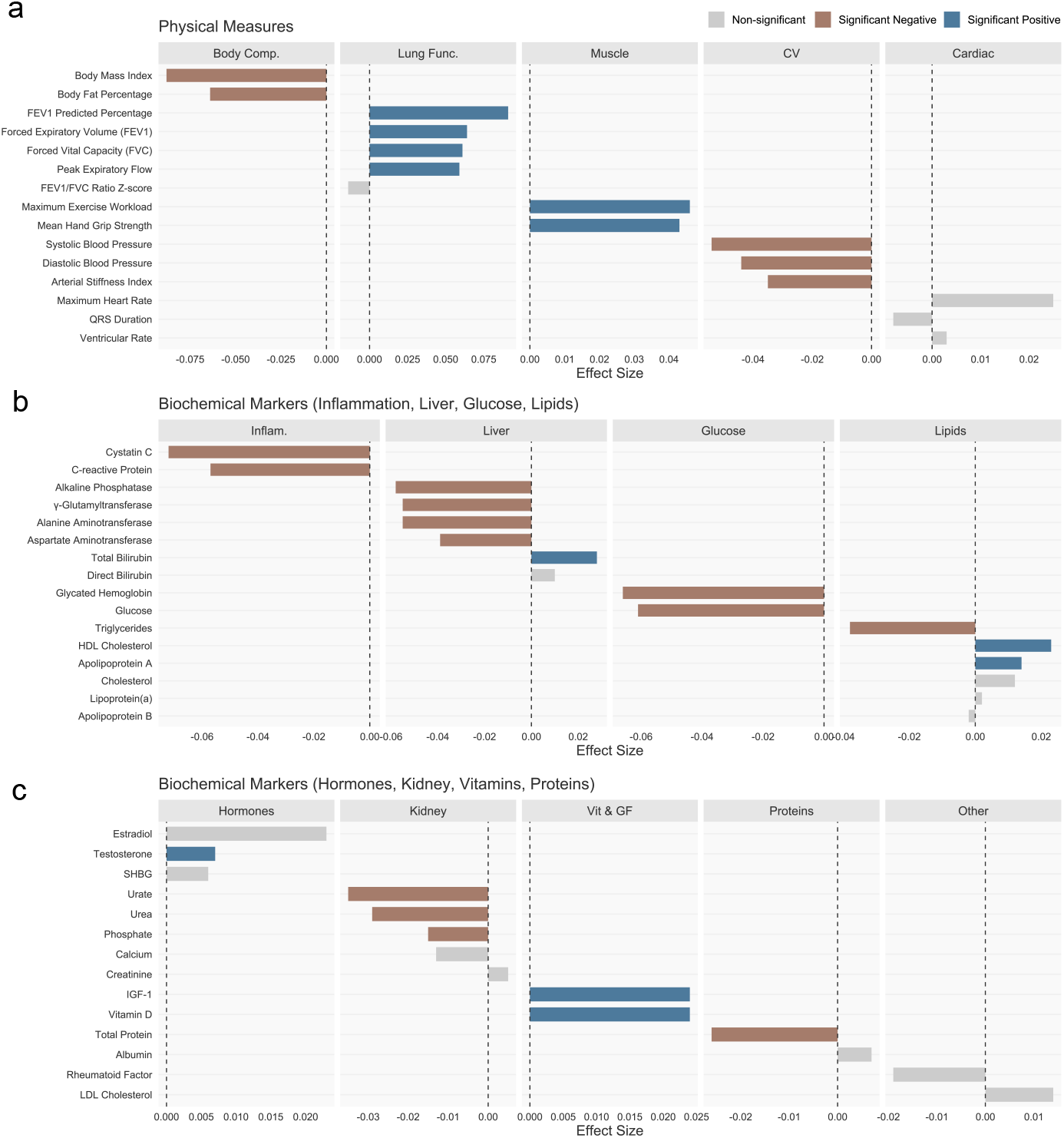
Long-term associations between childhood chickenpox infection and health parameters in adulthood. **a**. Physical measures include Body Composition (Body Comp.), Lung Function (Lung Func.), Muscular Strength (Muscle), Cardiovascular (CV), and Cardiac parameters; **b**. Metabolic markers include Inflammatory Markers (Inflam.), Liver Function (Liver), Glucose Metabolism (Glucose), and Lipid Metabolism (Lipids); **c**. Physiological markers include Hormonal Markers (Hormones), Kidney Function (Kidney), Vitamins & Growth Factors (Vit & GF), and Protein Markers (Proteins). Bar height represents standardized effect size adjusted for age, sex, socioeconomic status, and other childhood infections, with positive values indicating higher levels and negative values indicating lower levels in individuals with history of chickenpox infection. Colored bars indicate statistically significant associations (p < 0.05), with blue representing positive associations and red representing negative associations.

Among biochemical markers (**Fig. 2b**), Varicella infection showed significant inverse associations with inflammatory and metabolic markers, most strongly with cystatin C (*β* = −0.072, *p <* 0.001), glycated hemoglobin (*β* = −0.066, *p <* 0.001), glucose (*β* = −0.061, *p <* 0.001), and C-reactive protein (*β* = −0.057, *p <* 0.001). Consistent negative associations were observed with liver function tests, kidney function markers, and triglycerides, while positive associations were seen with HDL cholesterol, insulin-like growth factor 1, and vitamin D.

Analysis of blood count parameters ((**Extended Data Fig. 4**)) revealed negative associations between Varicella infection and neutrophil count (*β* = −0.067, *p <* 0.001), total white blood cell count (*β* = −0.055, *p <* 0.001), and various reticulocyte parameters. While neutrophil percentage was negatively associated (*β* = −0.032, *p <* 0.001), lymphocyte percentage showed a positive association (*β* = 0.030, *p <* 0.001), suggesting a shift in immune cell balance. These complex patterns suggest that prior Varicella infection may have subtle but long-lasting effects on hematopoiesis and immune cell distributions.

## Discussion

In this large-scale cohort study involving over 500,000 UK Biobank participants, we found that documented early-life Varicella infection was associated with significantly reduced risks of multiple chronic diseases in later life, with particularly strong protective associations for neurodegenerative conditions. These inverse associations were consistently stronger in women, increased with age, and remarkably, appeared to outweigh the well-established effects of socioeconomic deprivation on disease risk. Mendelian randomization analyses supported causality for several outcomes, strengthening confidence in our findings. Furthermore, prior Varicella infection was associated with favorable baseline profiles across multiple physiological systems, suggesting potential biological pathways through which early infection might shape long-term health trajectories.

Our findings present an intriguing paradox when considered alongside the growing literature on VZV and chronic disease. While we demonstrate protective associations between early-life Varicella infection and multiple chronic diseases, previous studies have shown that later-life VZV reactivation as herpes zoster is associated with increased risks of similar conditions [10]. This apparent contradiction may be reconciled by considering the temporal dimension of host-pathogen interactions across the lifespan.

Primary VZV infection typically occurs during childhood when the immune system is still developing. Our data show that the median age of Varicella infection was 7.0 years, decades before the onset of chronic diseases. At this developmental stage, the exposure may induce beneficial training of the innate immune system through mechanisms involving epigenetic reprogramming and metabolic rewiring of immune cells and their progenitors [8]. This process, termed “trained immunity,” can persist for months to years and involves substantial chromatin remodeling at promoters and enhancers of inflammatory genes [13].

Conversely, VZV reactivation as herpes zoster typically occurs in later life when immunosenescence is already underway. The inflammatory response to reactivation may exacerbate underlying pathological processes in individuals already at risk for chronic diseases. This interpretation is supported by recent findings that preventing herpes zoster through vaccination is associated with reduced dementia risk [11]. Together, these seemingly contradictory findings suggest that VZV influences health differently depending on the timing of exposure and the immunological context—beneficial training in early life versus harmful inflammation in later life.

The protective associations we observed between Varicella infection and chronic diseases may be explained by several mechanistic pathways. The concept of trained immunity provides a compelling framework [8]. Unlike classical immunological memory, which relies on the clonal expansion of antigen-specific T and B cells, trained immunity involves long-term functional reprogramming of innate immune cells through epigenetic modifications. This process can enhance responsiveness to subsequent immune challenges and alter inflammatory set points.

VZV infection may induce trained immunity at multiple cellular levels. In circulating monocytes and tissue macrophages (peripheral trained immunity), VZV exposure could leave an “epigenetic scar” at the level of stimulated genes, potentially altering their long-term responsiveness [14]. More critically, VZV might induce changes in bone marrow progenitors (central trained immunity), explaining the durability of protective effects over decades despite the short lifespan of mature myeloid cells [15]. Our findings of altered immune cell distributions in adults with prior Varicella infection, including lower neutrophil counts and higher lymphocyte percentages, support lasting effects on hematopoiesis.

The associations between Varicella infection and improved metabolic parameters (lower BMI, reduced glycated hemoglobin, lower glucose) are particularly noteworthy, as metabolic rewiring is essential for trained immunity [16]. Specifically, increased aerobic glycolysis, glutaminolysis, and cholesterol synthesis are hallmarks of trained immunity, with metabolites such as fumarate and mevalonate acting as key molecular mediators by modulating epigenetic enzymes [17]. The inverse associations we observed between Varicella infection and inflammatory markers (cystatin C, C-reactive protein) further suggest modulation of inflammatory pathways, potentially explaining protection against conditions with inflammatory components.

A striking aspect of our findings was the consistently stronger protective associations observed in women across all diseases. This sex-specific effect mirrors recent findings by Taquet et al. [11], who reported that the protective effect of recombinant shingles vaccine against dementia was 9% greater in women than men. These parallel observations from two different studies examining different aspects of VZV exposure strongly suggest that biological sex modifies the immune response to this virus.

Several mechanisms might explain these sex differences. Women generally mount stronger immune responses to infections and vaccinations, partly due to the immunomodulatory effects of sex hormones, particularly estrogens [18]. Estrogens enhance both innate and adaptive immune responses and may potentiate trained immunity processes. The X chromosome contains numerous immune-related genes, and incomplete X inactivation may result in higher expression of these genes in women [19]. Additionally, epigenetic reprogramming, central to trained immunity, may occur differently in men and women due to sex-specific chromatin landscapes [20].

Our findings suggest that sex should be considered not merely as a biological variable but as a fundamental modifier of host-pathogen interactions that shapes lifelong health trajectories. Future research into infectious exposures and chronic disease risk should systematically investigate sex-specific effects rather than simply adjusting for sex in statistical models.

Perhaps the most surprising finding was that the protective association of Varicella infection appeared to outweigh the well-established effects of socioeconomic deprivation on chronic disease risk. For Alzheimer’s disease, individuals with prior Varicella infection but high socioeconomic deprivation had lower risk than those without Varicella infection but favorable socioeconomic circumstances. This pattern challenges conventional understanding of health determinants, where socioeconomic factors are typically considered primary drivers of health disparities.

These findings raise intriguing questions about the interplay between biological and social determinants of health. While socioeconomic disadvantage typically increases exposure to health risks, certain childhood infections might paradoxically confer immunological advantages that partially mitigate these risks. This interpretation aligns with the “old friends hypothesis,” which posits that reduced exposure to microorganisms in more affluent environments may contribute to immune dysregulation and increased susceptibility to inflammatory conditions [7].

However, these results should not be interpreted as diminishing the importance of addressing socioeconomic health disparities. Rather, they highlight the complex, multifactorial nature of chronic disease etiology and suggest that interventions targeting socioeconomic factors alone may be insufficient without considering biological interactions with environmental exposures, including infections.

Our findings raise important considerations for varicella vaccination policies. Widespread childhood vaccination against varicella, implemented in many countries since the 1990s, has successfully reduced disease burden and complications from acute infection. However, the potential long-term immunological consequences of preventing natural infection merit careful consideration.

If natural VZV infection in childhood indeed confers protection against multiple chronic diseases through trained immunity, widespread vaccination could theoretically alter these protective effects. However, several factors complicate this interpretation. First, live attenuated varicella vaccines contain attenuated VZV and may themselves induce some degree of trained immunity, though likely differing from natural infection in magnitude and quality. Second, vaccination prevents herpes zoster in later life, which our findings and others suggest is harmful. The net effect of varicella vaccination on chronic disease risk thus represents a complex calculus of benefits and potential trade-offs across the lifespan.

Interestingly, a parallel emerges with recent findings regarding recombinant shingles vaccination in older adults, which is associated with reduced dementia risk [11]. Together, these observations suggest that both preventing early-life VZV infection through childhood vaccination and preventing later-life VZV reactivation through adult shingles vaccination might influence chronic disease risk, albeit through different mechanisms. This highlights the importance of life-course approaches to understanding how infections shape health trajectories.

Our study has several strengths, including its large sample size, prospective design, comprehensive assessment of health parameters, adjustment for multiple confounders, and use of Mendelian randomization to strengthen causal inference. However, several limitations warrant consideration.

First, ascertainment of prior Varicella infection relied on hospital records, which likely underestimate true exposure given that many cases are mild and treated at home. This misclassification would generally bias our results toward the null, suggesting our estimates may be conservative. Second, despite extensive adjustment, residual confounding by unmeasured factors cannot be ruled out in observational analyses, though our Mendelian randomization results mitigate this concern for several outcomes. Third, UK Biobank participants are generally healthier and more affluent than the broader UK population, potentially limiting generalizability. Finally, our study focused on a predominantly White European population, and findings may not apply to all ancestry groups due to potential differences in genetic susceptibility to VZV and chronic diseases.

Our findings open several avenues for future research. Mechanistic studies are needed to elucidate the precise molecular and cellular pathways through which early VZV infection might influence chronic disease risk decades later. Longitudinal studies with immunological assessments from childhood through adulthood could track the evolution of trained immunity phenotypes following natural infection versus vaccination. Similar investigations of other childhood infections would reveal whether the protective associations we observed are specific to VZV or represent a broader phenomenon.

From a public health perspective, our findings underscore the need for nuanced approaches to infectious disease prevention that consider potential trade-offs across the lifespan. Research comparing long-term health outcomes in birth cohorts before and after implementation of universal varicella vaccination could provide valuable insights into the population-level impacts of altering natural infection patterns.

Our study provides compelling evidence that early-life Varicella infection is associated with reduced risk of multiple chronic diseases in later life, with particularly strong protection against neurodegenerative conditions. These associations appear causal for several outcomes, are stronger in women, increase with age, and may partly mitigate socioeconomic health disparities. The findings challenge simplified views of infections as uniformly harmful and highlight the complex, time-dependent interactions between infectious exposures and chronic disease risk across the lifespan. Understanding how early-life infections shape health trajectories could inform more holistic approaches to disease prevention that consider both the immediate benefits of preventing acute infections and the potential long-term immunological consequences of altering natural exposure patterns.

## Methods

### Study design and data source

We used data from the UK Biobank, a large-scale biomedical database containing in-depth genetic and health information from approximately 500,000 UK participants recruited between 2006 and 2010 [21]. Participants were aged 40-69 years at recruitment and underwent comprehensive baseline assessments. Follow-up data were collected through linkage to national health registries until January 1, 2024. The study protocol was approved by the North West Multi-centre Research Ethics Committee, and all participants provided written informed consent. This study follows Strengthening the Reporting of Observational Studies in Epidemiology (STROBE) guidelines.

### Cohorts and exposures

Exposure to varicella-zoster virus (VZV) infection was determined through hospital records using ICD-10 code B01. Participants were classified as having prior VZV exposure if hospital records indicated a diagnosis of varicella before their baseline assessment date. We excluded participants with prevalent disease at baseline from the respective outcome analyses to prevent reverse causation.

### Covariates

We adjusted for multiple potential confounders including sociodemographic factors, comorbidities, and lifestyle variables. Sociodemographic factors included age at baseline, sex (female or male), ethnicity (White or non-White), household income (five categories: *<*$18,000, $18,000–30,999, $31,000–51,999, $52,000–100,000, and *>*$100,000), education level (college education: yes or no), and area-based socioeconomic deprivation measured by the Townsend Deprivation Index (TDI). For stratified analyses, the TDI was categorized into tertiles (low, medium, and high deprivation).

Lifestyle factors included smoking status (non-smoker or smoker, with the latter encompassing both current and former smokers), physical activity level (categorized as low, medium, or high based on combined moderate and vigorous activity scores, with thresholds at 600 and 1200 units), sleep duration (short: *<*7 hours, normal: 7-9 hours, or long: *>*9 hours), and body mass index (BMI). Health status indicators included hypertension (defined as systolic blood pressure *≥*140 mmHg or diastolic blood pressure *≥*90 mmHg).

### Outcomes

The primary outcomes were first diagnoses of six chronic diseases from baseline to end of follow-up in a time-to-event analysis: Alzheimer’s disease (AD: ICD-10 codes G30, F00), cardiovascular disease (CVD: I20-I25, I60, I61, I63, I64), chronic kidney disease (CKD: N18), chronic obstructive pulmonary disease (COPD: J44), Parkinson’s disease (PD: G20-G22), and type 2 diabetes (T2D: E11). For composite outcomes (AD, CVD, PD), we used the earliest date of any constituent diagnosis.

Secondary outcomes included physical measures (body composition, lung function, muscular strength, cardiovascular parameters) and biochemical markers (liver function, kidney function, glucose metabolism, lipid metabolism, inflammatory markers, hormones, and nutritional markers) measured at baseline assessment.

### Statistical analyses

We employed Cox proportional hazards models to estimate hazard ratios (HRs) and 95% confidence intervals (CIs) for the association between prior VZV infection and risk of chronic diseases. The proportional hazards assumption was tested using Schoenfeld residuals. Follow-up time was calculated from baseline assessment until diagnosis, death, or end of follow-up, whichever occurred first.

As a complementary approach, we used logistic regression models to estimate odds ratios (ORs) for the association between VZV exposure and disease outcomes, adjusting for the same covariates as in the Cox models. Stratified analyses were conducted by age group (*≤*55, 55-64, and 65-70 years), sex, and socioeconomic deprivation tertiles to investigate potential effect modification. Interaction terms were included in the models to test for significant differences between subgroups.

For secondary outcomes consisting of continuous health parameters, we employed linear regression models with standardized outcome variables (z-scores) to facilitate comparison across different metrics. All continuous outcomes were standardized (mean=0, SD=1) before analysis, with models adjusted for the same set of covariates as in the primary analyses.

To strengthen the causal inference, we utilized a two-sample Mendelian randomization (MR) approach using summary-level data from genome-wide association studies (GWAS) conducted on individuals of European ancestry. The exposure of MR analysis was the endpoint of varicella, with instrumental variables derived from the FinnGen GWAS. For the outcomes of AD, CVD, CKD, COPD, PD, and T2D, instrumental variables were identified from independent GWASs from UKB and European Bioinformatics Institute (EBI). In MR analysis, we used odds ratios (OR) and 95% CI to estimate the disease risk caused by varicella history. For the primary MR analysis, we utilized the random-effects inverse-variance weighted (RE-IVW) method to estimate the causal effect. Several other MR analyses, including IVW, fixed effects IVW, MR-Egger regression, and weighted median (WM) were performed to assess the robustness of the results. We used the MR-Egger intercept test to assess for horizontal pleiotropy and Cochran’s Q statistic for population heterogeneity. To confirm the causal effect of any single SNP, we conducted a leave-one-out (LOO) analysis by discarding each exposure-associated SNP and repeatedly performing IVW analysis. We also performed a bidirectional two-sample Mendelian randomization analysis from the reverse direction to examine potential reverse causality.

Missing data were handled using complete case analysis. For each outcome analysis, we excluded participants with missing data on the exposure, outcome, or any of the covariates included in the fully adjusted model. All statistical tests were two-sided, with p-values < 0.05 considered statistically significant. Statistical analyses were conducted in R version 4.1.0.

## Data availability

UK Biobank data are publicly available at https://www.ukbiobank.ac.uk/ (application number 146760).

## Acknowledgements

This work was supported by the Singapore Ministry of Education Tier 2 grant A-8001562-00-00 and the Tier 1 grant A-8002931-00-00 at the National University of Singapore.

---

**Extended Data Table 1.**
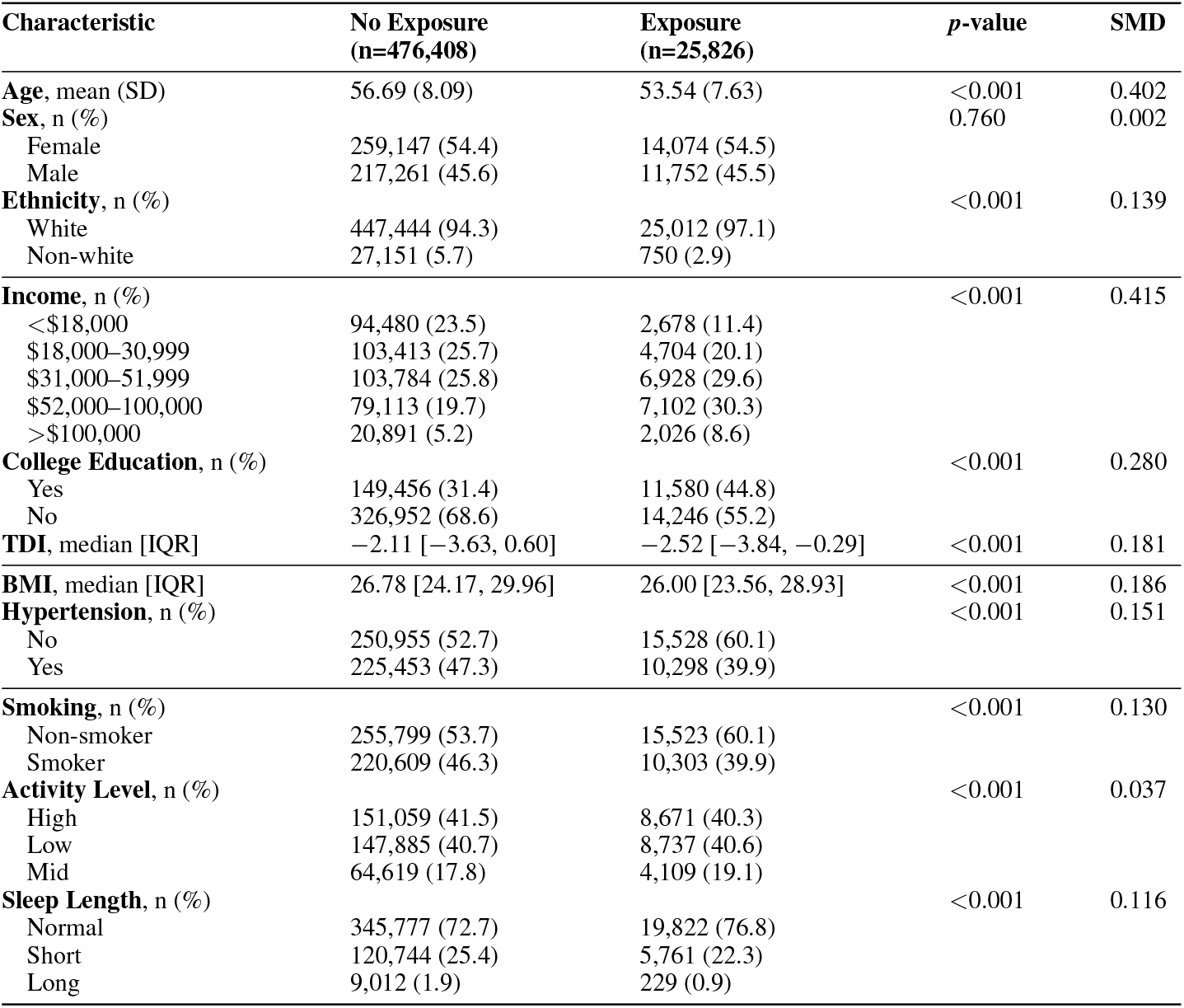
Baseline Characteristics by Varicella Exposure Status. SMD = Standardized Mean Difference; TDI = Townsend Deprivation Index; IQR = Interquartile Range; SD = Standard Deviation.

**Extended Data Fig. 1.**
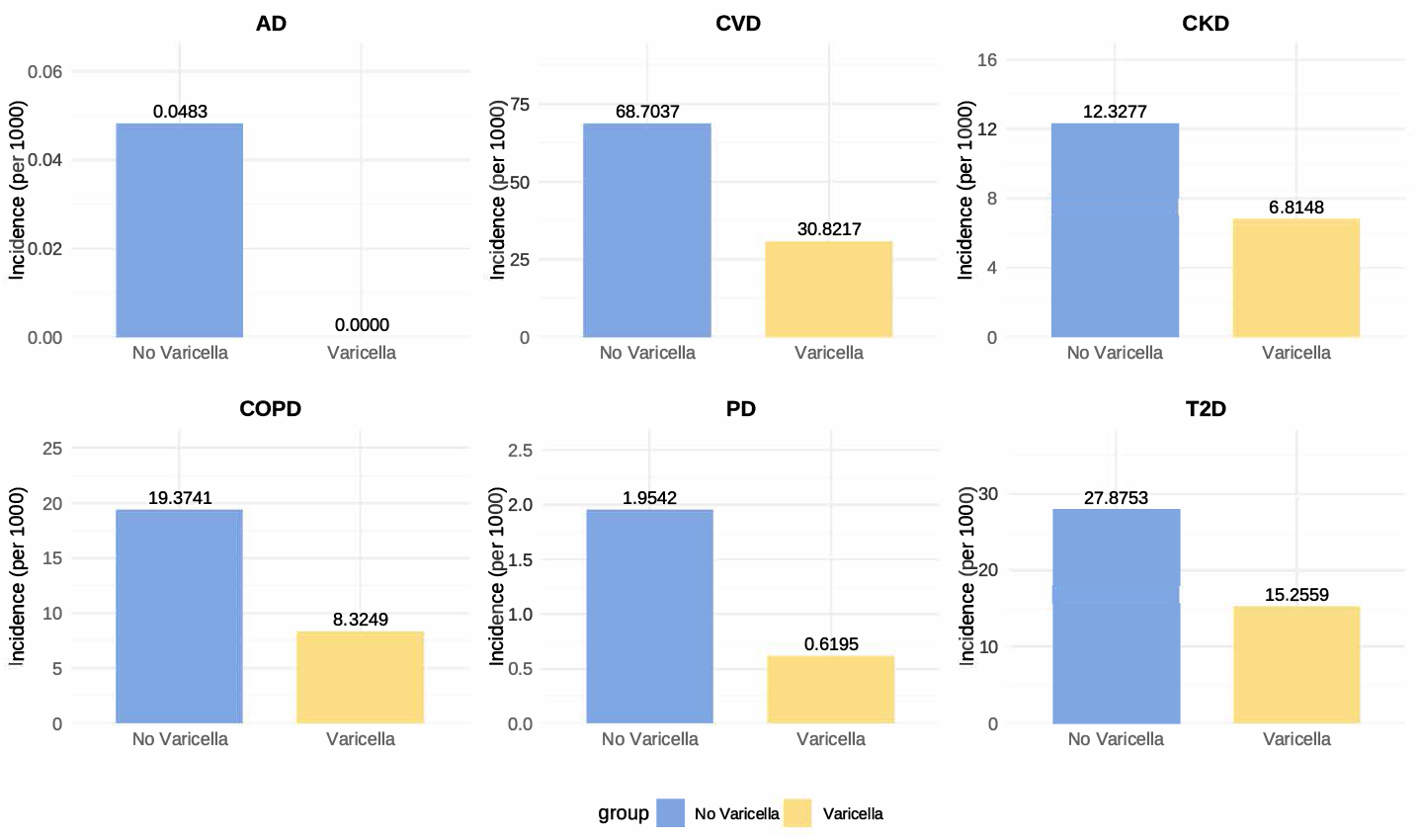
Comparison of baseline disease incidence rates between individuals with prior varicella infection (Varicella) and those without (No Varicella) in the UK Biobank cohort.

**Extended Data Fig. 2.**
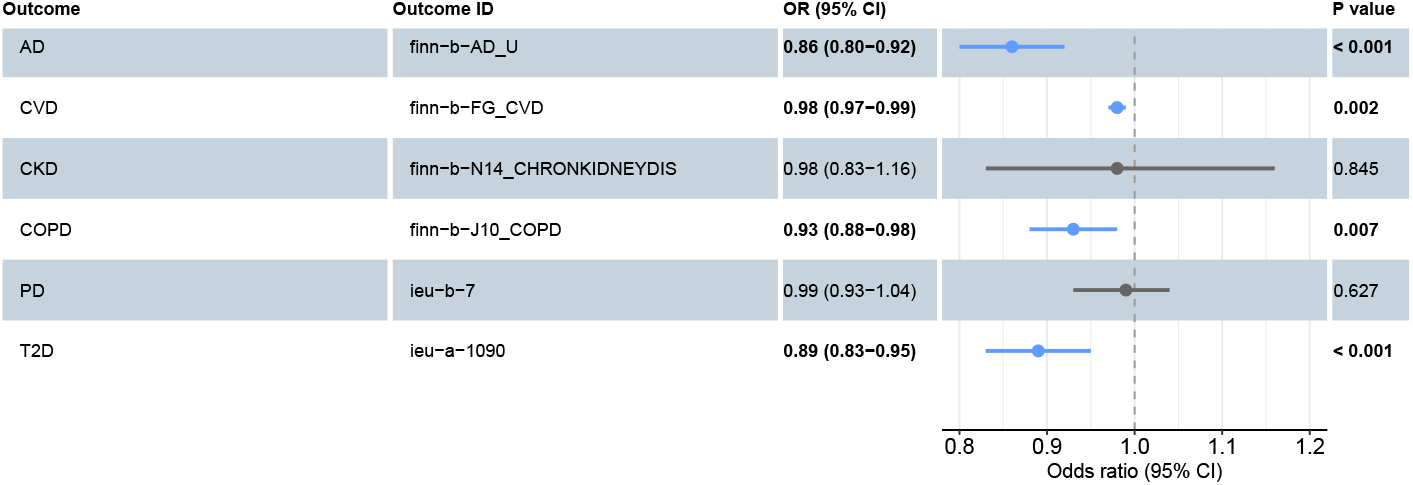
Mendelian Randomization analysis showing the causal effects of varicella zoster infection on the risk of six common diseases.

**Extended Data Fig. 3.**
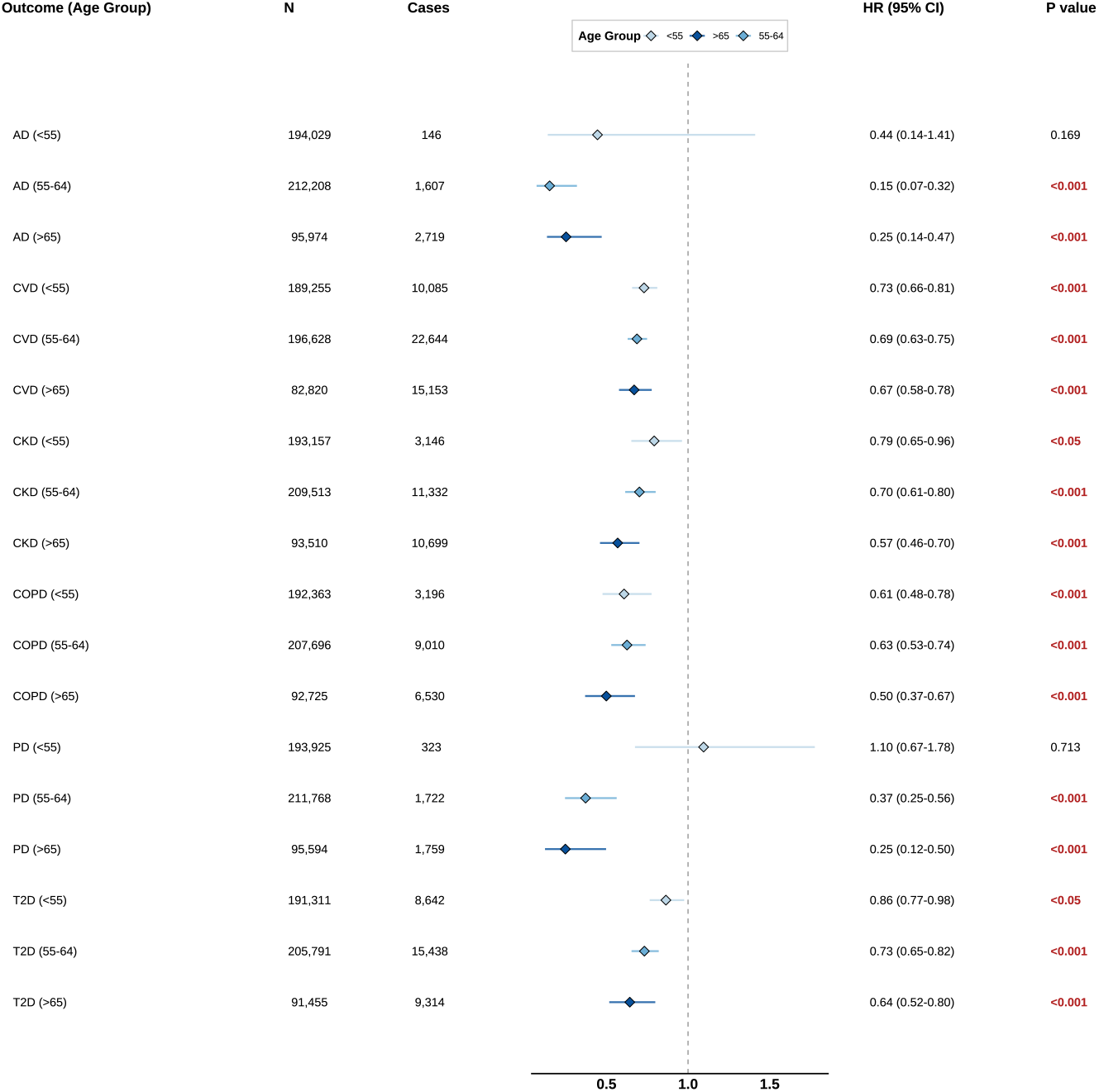
Association between prior varicella infection and chronic disease risk, stratified by age group. The forest plot displays hazard ratios with 95% confidence intervals from Cox proportional hazards models. Each model was adjusted for sex, ethnicity, income, education, Townsend deprivation index, smoking status, sleep duration, BMI, and physical activity. Age stratification was performed using three groups: *≤*55, 55-64, and 65-70 years. Bold red p-values indicate statistical significance (p < 0.05).

**Extended Data Fig. 4.**
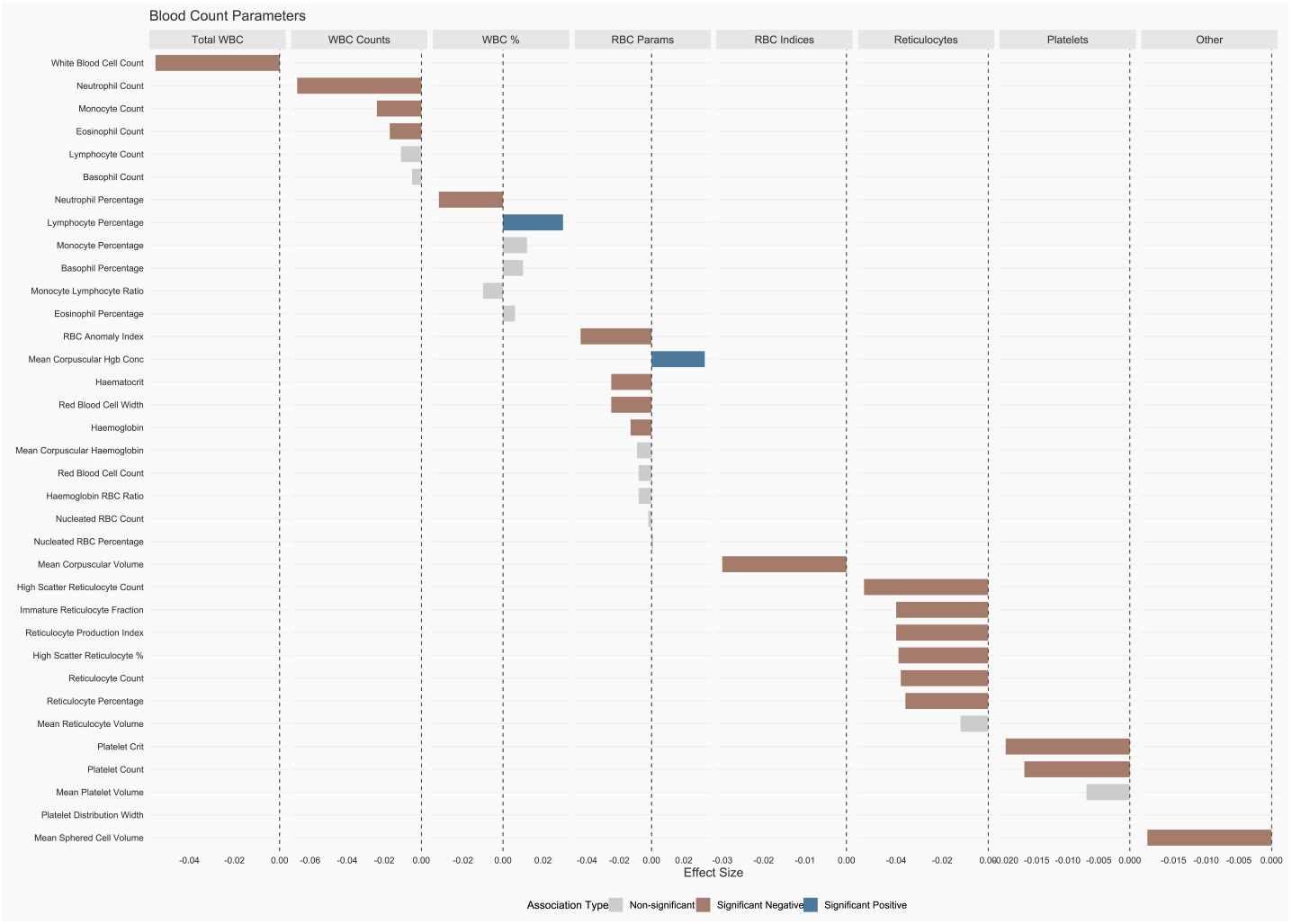
Association between chickenpox infection and blood count parameters. This figure depicts standardized effect sizes for the relationship between chickenpox infection and various hematological parameters. Each bar represents a specific blood parameter, with bar height indicating the magnitude of the effect size; positive values indicate that chickenpox infection is associated with higher parameter values, while negative values indicate association with lower values. Color coding: dark blue indicates significant positive associations (p < 0.05), brown indicates significant negative associations (p < 0.05), and gray indicates non-significant associations (p *≥*0.05). Blood parameters are grouped by functional categories: Total White Blood Cell Count (Total WBC), White Blood Cell Differential Counts (WBC Counts), White Blood Cell Percentages (WBC %), Red Blood Cell Basic Parameters (RBC Params), Red Blood Cell Indices (RBC Indices), Reticulocyte Parameters (Reticulocytes), and Platelet Parameters (Platelets). Within each category, parameters are ordered by absolute effect size magnitude in descending order.

## Notes

### Competing Interest Statement

The authors have declared no competing interest.

### Author Declarations

UK Biobank data are publicly available at https://www.ukbiobank.ac.uk/ (application number 146760).

